# The Heart Hive: direct-to-participant recruitment provides an effective, scalable route to identify engaged participants for cardiomyopathy research

**DOI:** 10.64898/2025.12.03.25339687

**Authors:** Angharad M Roberts, Rachel J Buchan, Pantazis Theotokis, George Powell, Lara Curran, Daniel J Hammersley, Matthew Edwards, Emma Jennings, Esme Cavanagh, Rachel Cartwright, Upasana Tayal, Brian P Halliday, Sanjay K Prasad, James S Ware

**Affiliations:** Imperial College London; National Heart and Lung Institute, Imperial College London; MRC Laboratory of Medical Sciences, Imperial College London; British Heart Foundation Centre of Research Excellence, School of Cardiovascular and Metabolic Medicine and Sciences King’s College London; Royal Brompton and Harefield Hospitals, Guys and St Thomas NHS Foundation Trust

**Author notes:** These authors contributed equally (Angharad M Roberts, Rachel J Buchan).

**Keywords:** Cardiomyopathy, Direct-to-patient, Patient–study matching, Patient-reported outcomes (PROs), Remote recruitment

## Abstract

**Background:** Direct-to-patient platforms can overcome geographic and capacity barriers to rare-disease research, supporting inclusive and scalable recruitment. The Heart Hive was developed to enable nationwide, genomics-integrated cardiomyopathy research and rapid patient–study matching through a digital, patient-centred platform.

**Methods:** Adults self-reporting a diagnosis, or inherited risk, of cardiomyopathy enrolled online, completed electronic consent, and provided baseline and longitudinal data for study matching. Participants in the pilot Cardiomyopathy Study were invited to return saliva kits by post for central cardiomyopathy-panel sequencing. Engagement and platform development were supported through formal patient and public involvement and partnership with Cardiomyopathy UK. Feasibility of digital recruitment, retention, and genomic validation was assessed.

**Results:** 338 participants met eligibility; 227 (67%) completed baseline surveys, 195 (86%) returned saliva kits, and 174 were sequenced (98 DCM, 76 HCM). Follow-up survey completion among sequenced participants was 83%. The cohort was 57% female, median age 57.5 years, and 98% White European; 78% had never previously participated in cardiovascular research. Genetic architecture was comparable to a clinic-recruited cohort (DCM 34.7% vs 30.6%; HCM 44.7% vs 38.2%), and actionable variants were identified in 17% of DCM and 29% of HCM participants opting for personal results.

**Conclusions:** Self-reported recruitment accurately identifies cardiomyopathy cohorts equivalent in genetic composition to clinic-ascertained populations. The Heart Hive demonstrates a decentralised, re-contactable, and registry-ready digital model for nationwide precision-cardiology research.

## Introduction

Cardiomyopathies are primary disorders of heart muscle, which often have a genetic aetiology, and represent an important health burden. In the western world more than 1 in 250 people are affected. Dilated cardiomyopathy (DCM) and hypertrophic cardiomyopathy (HCM) are the most common types and are leading causes of sudden death and heart transplantation. While a monogenic cause is identified in an important proportion of cases, primarily due to variants in genes encoding components of the sarcomere, the majority remain genetically unexplained. The high variability in penetrance and expressivity of cardiomyopathies makes variant interpretation and clinical prognostication challenging. There is an important need for research to understand the genetic architecture of cardiomyopathies, to characterise their natural history, to understand the interplay of genetic and environmental modifiers of disease, and to develop biomarkers for diagnosis, prognostication, and therapeutic stratification.

Contemporary genetics, particularly for complex conditions like cardiomyopathy requires large cohorts for well-powered studies; these are beyond the reach of single centres and even stretch traditional collaborative networks. Researchers face major challenges in gathering adequately sized cohorts and in maintaining patient engagement. Recruitment is often centred around larger teaching hospitals and specialist clinics that may not represent the full phenotype spectrum of disease, and other disparities in accessing healthcare. This recruitment bias also leaves research deserts where patients struggle to access research studies and trials. Highly stratified approaches often require a set of very specific characteristics which would benefit from having a large pool of participants from which to identify rare subsets such as cardiomyopathy caused by specific genotypes. For longitudinal studies of risk factors and outcomes, ongoing engagement is key. Various approaches have been applied to maximise recruitment, typically at the cost of engagement and study completion. The legacy of the COVID-19 pandemic has added further difficulties to research recruitment, with more patients attending virtual clinics and reducing in-person hospital visits, limiting opportunities to discuss research with patients directly.

To address these challenges, direct-to-patient recruitment approaches offer a way to engage with patients who wish to take part in research, provide a route for participants to connect with researchers, and develop a community of research participants. Enabling patients to self-identify as research-willing, and to proactively select research studies, is a powerful driver toward both recruitment and engagement.

### Can online recruitment help researchers to access cardiomyopathy patients who have previously been difficult to recruit into studies and are they representative of the patient population?

To address these problems, we set up the Heart Hive, an online platform to connect researchers to potential research participants and build a registry of research-willing people with cardiomyopathy and myocarditis. In this paper we report the results of the pilot study and the feasibility of direct to participant recruitment for cardiomyopathy.

## Methods

The Heart Hive (www.thehearthive.org) was established in late 2019 after close work with patient groups and Cardiomyopathy UK, a patient charity. At patient engagement events patients reported wanting to take part in more research and expressed frustration at being unable to identify research opportunities. The Heart Hive portal is open to people aged 18 and over with cardiomyopathy or myocarditis, a family history of cardiomyopathy, or a genetic variant associated with cardiomyopathy. Participants opt-in to be matched with and notified about new research or clinical trial opportunities, can decide which studies to enrol in, and whether to share their data with specific researchers.

The Heart Hive can be used to advertise trials to eligible participants, eConsent participants into studies, collect healthcare and outcome data with surveys, and collect saliva samples via the mail for genetic analysis. The Heart Hive Cardiomyopathy Study launched as a pilot study alongside the Heart Hive portal and aims to compare the DCM and HCM cohorts recruited online through the Heart Hive with traditional patient cohorts recruited via hospital clinics, to validate this route for participant identification and understand any limitations in online approaches.

### The Heart Hive Registry

The Heart Hive registry was developed as a web portal with secure GDPR compliant back-end database and facility to automate participant contact (e.g. send invite or reminder emails) and to present tailored options on a participant dashboard, with eligibility for opportunities determined by data provided during registration. In the UK, a registry and database do not require ethics (REC/IRB) approval, but any data use for research requires an ethically approved study. Participants provide permission to be contacted for future research invitation and marketing, which can be withdrawn as participants wish in the future.

### Registry technical capability

The Heart Hive enables a range of activities. It can be used to match participants with relevant study adverts, screen for study-specific eligibility criteria with questionnaires, provide study information and obtain informed consent, conduct study activity with patient reported outcome measures (PROMs), track study journey, send participant reminders, manage biosample collection, and communicate with participants directly. eConsent processes integrate participant information sheets (PIS) and consent forms, with check points to allow researchers to confirm participants’ understanding as they progress through the form. Surveys are automated and can be sent as one-off and recurring items at specified time-points to collect baseline and follow-up health information. Signed consent forms, PIS, and forms to withdraw from research can all be accessed by participants in their personal dashboard.

At enrolment participants self-report their diagnosis and provide some key demographic information. Using registration information, participants are selected for invitation to research studies that they may be eligible to take part in.

### Pre-launch/connecting with potential participants

In the lead up to launching the Heart Hive, we ran a crowdfunding campaign in collaboration with Crowdfunder (https://www.crowdfunder.co.uk/p/thehearthive) to raise both funds and awareness. Donations were matched by Cardiomyopathy UK. A focussed social media campaign was conducted on Facebook, Twitter (now X), and Instagram, in addition to blogs on our web pages and articles in Cardiomyopathy UK’s quarterly magazine ‘MyLife’ to raise awareness (Figure 1A).

**Fig 1.**
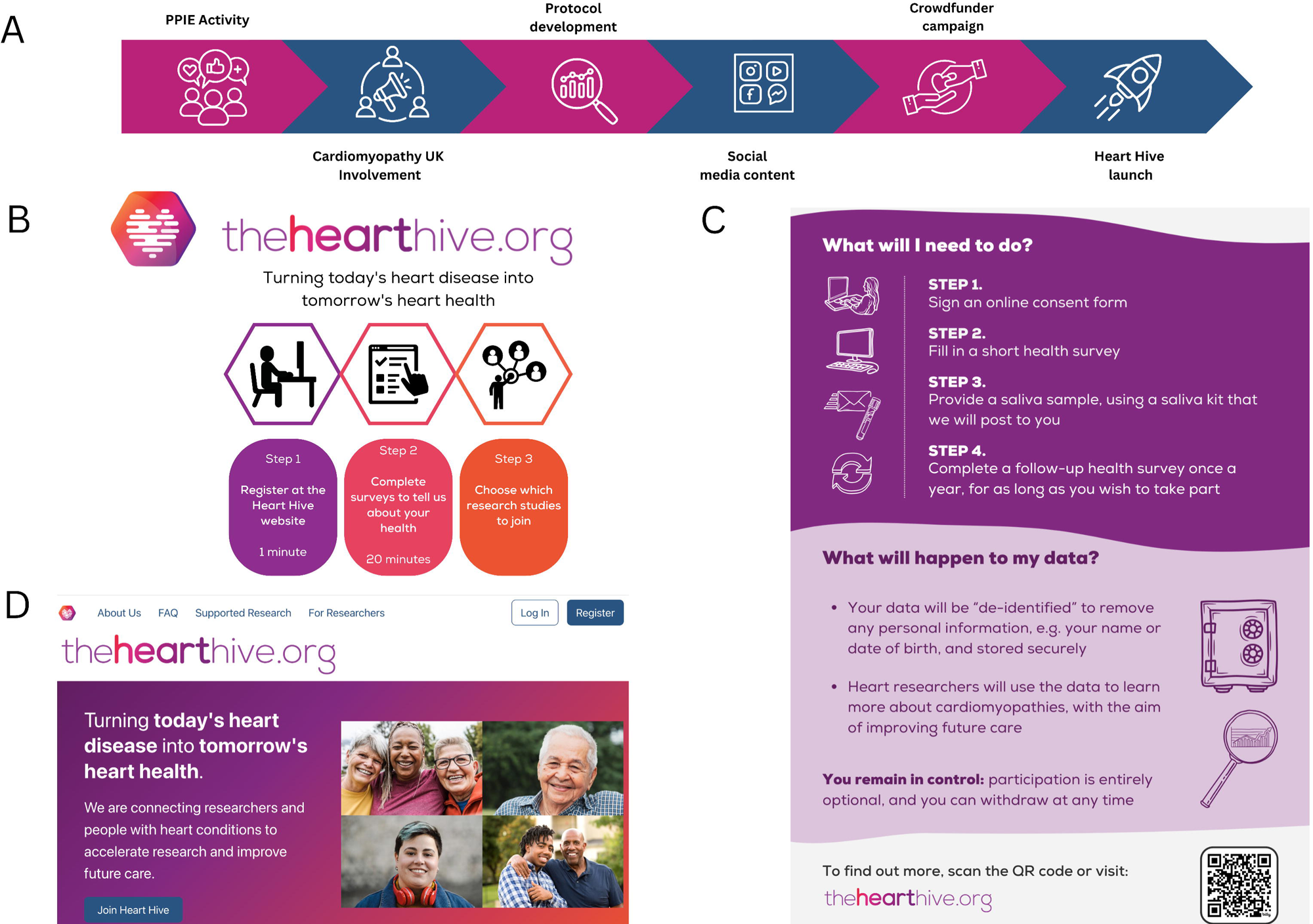
A. Timeline of Heart Hive development, crowdfunding, participant and patient engagement activities before launch in November 2019. B. Infographics used to explain the Heart Hive concept to potential participants C. Infographic detailing the Cardiomyopathy Study to potential participants D. The Heart Hive website frontpage – thehearthive.org

The web portal was originally developed with DatStat Inc., a commercial health care and clinical trials software provider. The platform was subsequently re-launched in collaboration the Broad Clinical Labs (a subsidiary of the Broad Institute) on the *Juniper* data donation platform (Figure 1D). During the initial stages of development, we collaborated closely with Cardiomyopathy UK and their members to ensure the content was relevant and easily accessible. We undertook extensive user experience testing with stakeholders representing a broad age-range, and from diverse ancestries, geographies, and socioeconomic groups.

### Baseline and Follow Up Surveys

All Heart Hive participants with DCM or HCM were invited to complete baseline health surveys tailored to their diagnosis (Figure 1B). The baseline survey collects details including cardiomyopathy diagnosis, family history, genetic testing, medication history, cardiovascular events, and demographics. We asked about outcomes as an indicator of disease severity: whether participants had experienced atrial fibrillation, strokes, cardiac arrests, had implanted devices, transplants (or were on the transplant list), left-ventricular assist devices or had myectomies.

Clinic recruited cohorts are screened for eligibility before recruitment, particularly for primary cardiomyopathy studies, including assessment of potential confounders or secondary causes. This information is typically gleaned from medical records before approaching the patient. We collected participant-reported data relating to comorbidities and confounders such as valve disease, diabetes, cholesterol, pregnancy history, chemotherapy, hypertension, other environmental causes of cardiomyopathy. We asked several questions as indicators of coronary artery disease (see box 1) and aggregated answers to assess possibility of ischaemic CM.

Participants are asked to complete a shorter survey annually, collecting details of any further events experienced, changes in health, medication, and treatment.

### The Heart Hive Cardiomyopathy Study

The Heart Hive’s first study was a validation and feasibility study to examine the recruitment, demographic trends and biases, and genetic architecture of DCM and HCM cohorts recruited through the Heart Hive platform.

Participants were invited to enrol in the Heart Hive Cardiomyopathy Study through the platform if they were age 18 or over, resident in the UK, and reported having DCM or HCM. The study invitation was presented on personalised dashboards after registration. Participants click through to a combined information sheet and eConsent form and are asked to complete the baseline survey tailored to their reported diagnosis if they have not already done so. Once both eConsent and baseline health survey were completed a saliva kit was sent out by post to collect a DNA sample (Figure 1C & Figure 2).

**Fig 2.**
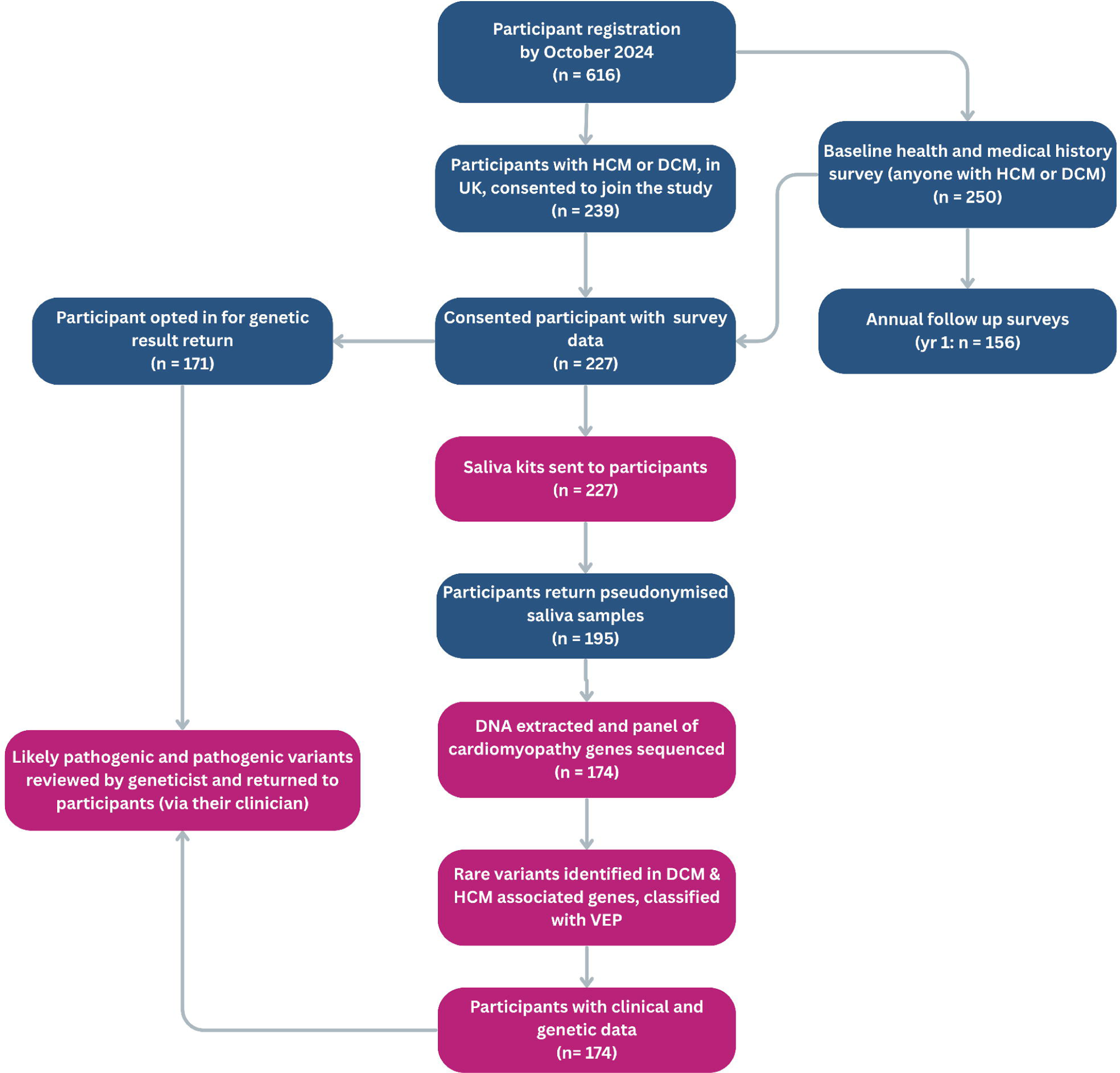
Chart describing study flow and user journey.

During consent, participants chose whether or not they wished to receive research genetics results that could be relevant to their cardiomyopathy diagnosis. For those who opted in, results are sent to the participants’ specialist cardiologist or geneticist (not directly to the patient). The study does not seek or report variants that are unrelated to cardiomyopathy.

The health surveys were carefully designed to capture information that would usually be screened in clinic when assessing eligibility criteria for studies, or when assessing adverse cardiovascular events in natural history studies or interventional trials (Box 1). Some eligibility criteria relate to capacity to give informed consent, and we address this by only offering the study to participants 18yrs and over, and by requiring that they are able to complete the eConsent and enter their own health data, both of which are tasks that demand an understanding of English and of their diagnosis.

### DNA Collection and sequencing

Participants were sent saliva collection kits (Norgen Biotek) through the mail and asked to provide a 1 ml sample of saliva. Return postage was provided and participants were asked to return to the study team for DNA extraction. DNA was extracted using the Qiasymphony DSP system (Qiagen) and quantified with Qubit BR dsDNA (Invitrogen). Targeted sequencing was performed with a 169 gene panel sequenced (TruSightCardio, Illumina) on the NextSeq 550 with 150 bp paired-end reads. Reads were mapped and annotated with a custom bioinformatic pipeline [1].

### Genetic architecture

We examined 11 genes associated with DCM and 8 sarcomeric genes associated with HCM [2]. Variants with a gnomAD population maximum allele frequency >= 0.001 were excluded. Variants were prioritised for burden testing if their predicted consequence had established evidence of pathogenicity in the relevant gene. For example, pathogenic variants in MYBPC3 act through haploinsufficiency, so protein-truncating variants were included. In contrast, MYH7 is not haploinsufficient but causes disease through altered gene products, so inframe deletions and missense variants were included. Protein truncating variants (PTV) in TTN were further filtered to exclude those in exons with PSI (percentage spliced in) < 90 % in the heart. Variants identified in Heart Hive and clinic recruited cohorts were identified and annotated in the same way. Differences between the Heart Hive and our clinic-recruited cohorts were assessed using Fisher’s exact tests. Multiple testing was addressed by applying a Bonferroni p-value correction. Variants for return to participants as relevant to their diagnosis were further manually assessed according to ACMG guidelines.

### Return of genetic results

Variants in genes relevant to the cardiomyopathy subtype were annotated with variant effect predictor [3] and classified according to the ACMG/AMP guidelines. Research results were reviewed by a UK accredited clinical scientist to confirm classification. Likely pathogenic and pathogenic variants in known DCM and HCM associated genes were reported back to the clinical teams of participants who had opted in to receive results.

### Comparison Cohorts

The Heart Hive cohort was compared with a cohort of cardiomyopathy patients recruited more traditionally at the Royal Brompton Hospital (RBH) Cardiovascular Biobank in London, UK (19/SC/0257), with matched sequencing (Illumina TruSight panel) and bioinformatic pipeline. The clinic-recruited cohort comprises 863 individuals with DCM and 381 individuals with HCM recruited between 2009-2016 and sequenced between 2015 and 2021.

To contextualise the demographic and behavioural characteristics of the Heart Hive cohort, we also compared our aggregated data to publicly available national health statistics from across the UK. These included: The Health Survey for England (HSE); The Scottish Health Survey (SHeS) for Scotland; The National Survey for Wales (NSW); Aggregated Health survey data in BHF Heart & Circulatory Disease Statistics 2024; NHS Digital Statistics on Public Health, England 2023.

Data were drawn from the most recent available survey cycles, and included variables on age, gender, education, smoking, and alcohol consumption. While methodological differences exist between surveys, they provide a broadly comparable baseline for evaluating representativeness of our UK-wide cohort. Where appropriate, ranges or weighted averages were used to approximate UK-wide norms.

### Ethics approval

The Heart Hive Cardiomyopathy study was approved by Health and Social Care Research Ethics Committee B (HSC REC B) reference 19/NI/0168 and the Royal Brompton & Harefield Cardiovascular Research Centre Biobank was approved by the South Central - Hampshire B Research Ethics Committee reference 19/SC/0257. All participants provided informed consent.

## Results

### Portal Sign Ups, Study enrolment

Over 900 participants with cardiomyopathy or myocarditis, or at risk of cardiomyopathy have signed up to the Heart Hive registry since launch. Thirteen studies have been offered to participants and several opportunities to join focus groups and patient perspective surveys. The first phase of CM study from launch to October 2024 was restricted to self-reported DCM and HCM, 338 UK participants were eligible to participate. Of these 239/338 (68 %) enrolled and 227 completed the baseline health survey. Saliva kits were sent to all 227 enrolees with baseline data and 195 (86 %) were returned (Figure 2).

Genetic data from the first tranche of samples to be sequenced (n = 174, received by August 2021) were analysed for the Heart Hive Cardiomyopathy pilot phase (HH-CM pilot) of the study. Follow-up surveys were collected from 83 % of sequenced participants.

### Cohort description

Full data for comparison for the Heart Hive cohorts, clinic recruited cohorts and general population is collected in supplementary table 1.

The HH-CM pilot cohort consisted of 98 DCM and 76 HCM participants, 57% female (61 % DCM, 53 % HCM), 98% white European ancestry with a median age of 57.5 years at recruitment (DCM 56.5, HCM 58.5), and 49 years at reported disease onset (HCM 48.5, DCM 49) (Figure 3 & supplementary table 1). Participants were recruited from across the UK, dispersed widely, not just restricted to larger urban centres (Figure 4). 78 % of participants in the HH-CM pilot had never previously taken part in cardiovascular research. Our participants recorded the name of their cardiologist and the centre they attend. For 56 % of the cohort their cardiology care was delivered by an Inherited Cardiac Condition (ICC) specialist service. 38 % (66/174) of participants reported a family history of cardiomyopathy (21 % did not know their family history status), and 59 % (103/174) reported having had genetic testing at registration, or in their year one or two follow up survey. 67 % (71/103) of those seen at an ICC service had genetic testing vs 46 % (32/71) at non-ICC centres *(p = 0.0027* chi-squared)

**Fig 3.**
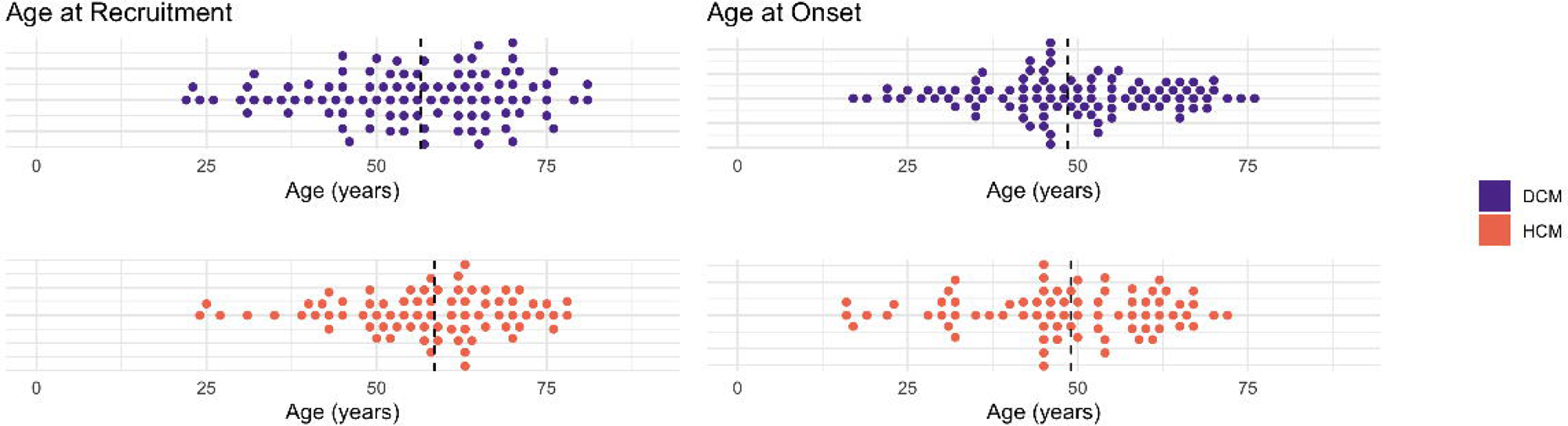
Participant age at recruitment and self-reported age of cardiomyopathy onset in the Heart Hive Cardiomyopathy Study. Dotted line represents median age.

**Fig 4.**
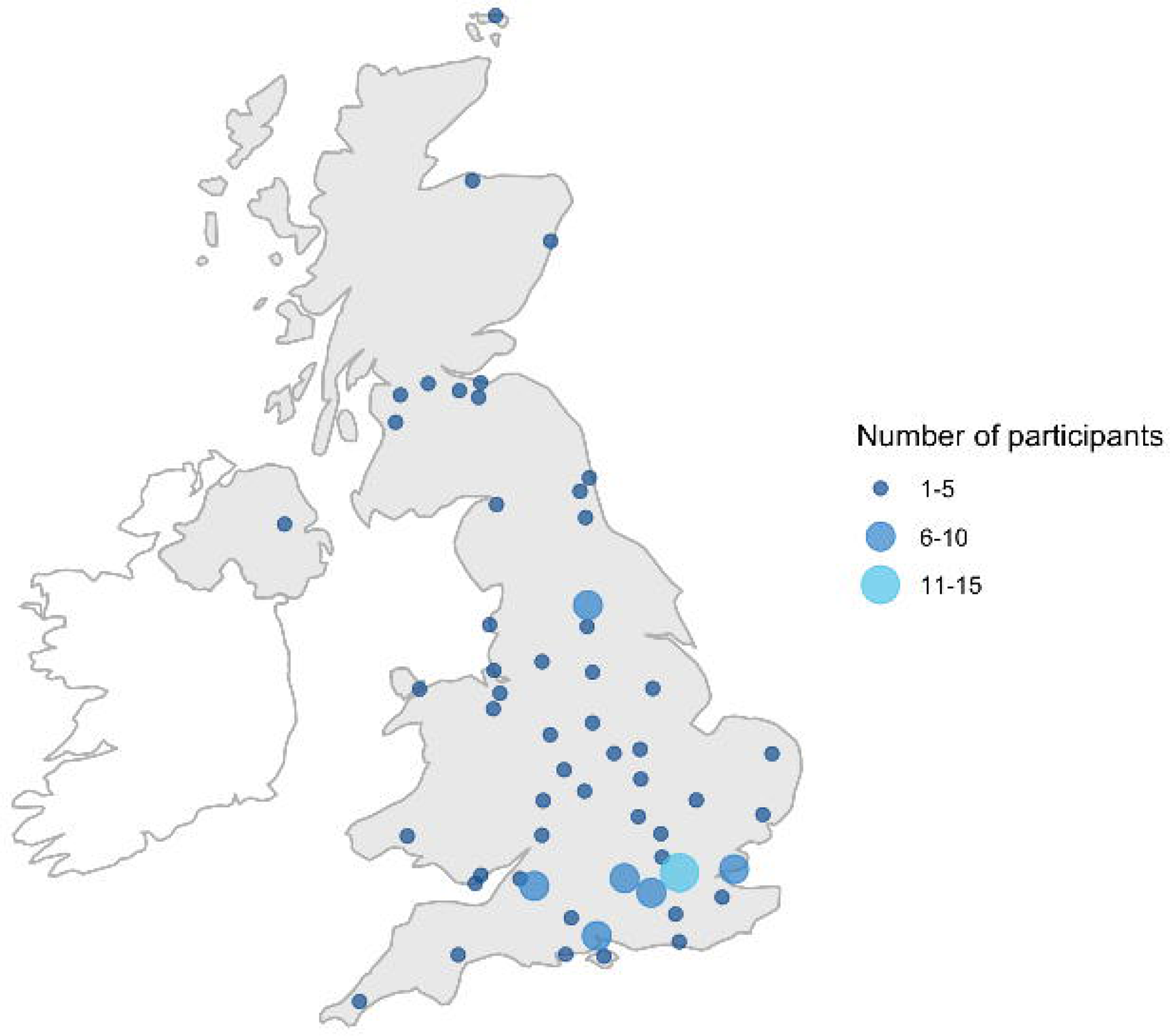
Geographical spread of Heart Hive Cardiomyopathy participants, plotted to nearest UK local administrative centres.

### Demographic comparison with UK general population from census & NHS Digital data

The HH-CM pilot cohort is 57 % female while the UK population is 51 % female. White ethnicity is reported by 98 % of HH-CM pilot participants and by 81-94 % of the UK (England, Wales, and Scotland) population. In terms of educational attainment, 85% of the HH-CM pilot cohort report having A-levels or higher, while in the UK population the figure is 41-44%. 74% of HH-CM pilot participant are married or in a domestic partnership, and 25% reported being either single, divorced, separated, or widowed; corresponding UK figures show 40% living in a couple household and 60% not living as a couple (includes living alone, single parent households and house shares). (See supplementary table 1) Only 1% our HH-CM pilot participants are current smokers and 32 % are ex-smokers, whereas 13-15 % of adults in the UK population report being smokers. 65% of HH-CM pilot participants currently drink alcohol and 30 % are former alcohol drinkers, while in the UK population 80-83% have drunk alcohol in the last 12 months. 5 % of our cohort have never drunk alcohol (11 % in the general population). (Figure 5 & supplementary table 1) 9% of HCM and 3% of DCM participants in the HH-CM pilot report current hypertension, and 47% and 23% controlled hypertension, compared to estimates of 15% hypertension in the general population. In the UK, the prevalence of raised cholesterol was 53 % (assessed by blood sample) compared to the HH-CM pilot cohort in which 9 % reported high cholesterol levels and a further 17 % reported a history of high cholesterol which was controlled. (Figure 5& supplementary table 1) 2.1 % of the UK population have AF, with a corresponding number of 51 % of DCM and 46 % of HCM participants in the HH-CM pilot. 1.9 % of the UK adults have had a transient ischemic attack (TIA) or cerebrovascular accident (CVA) whereas 2 % (DCM) and 5 % (HCM) of the HH-CM pilot report a TIA or CVA. 3.1 % of UK adults have coronary heart disease with a corresponding number of 11 % (DCM) and 9 % (HCM) of the HH-CM pilot group. (See supplementary table 1)

**Fig 5.**
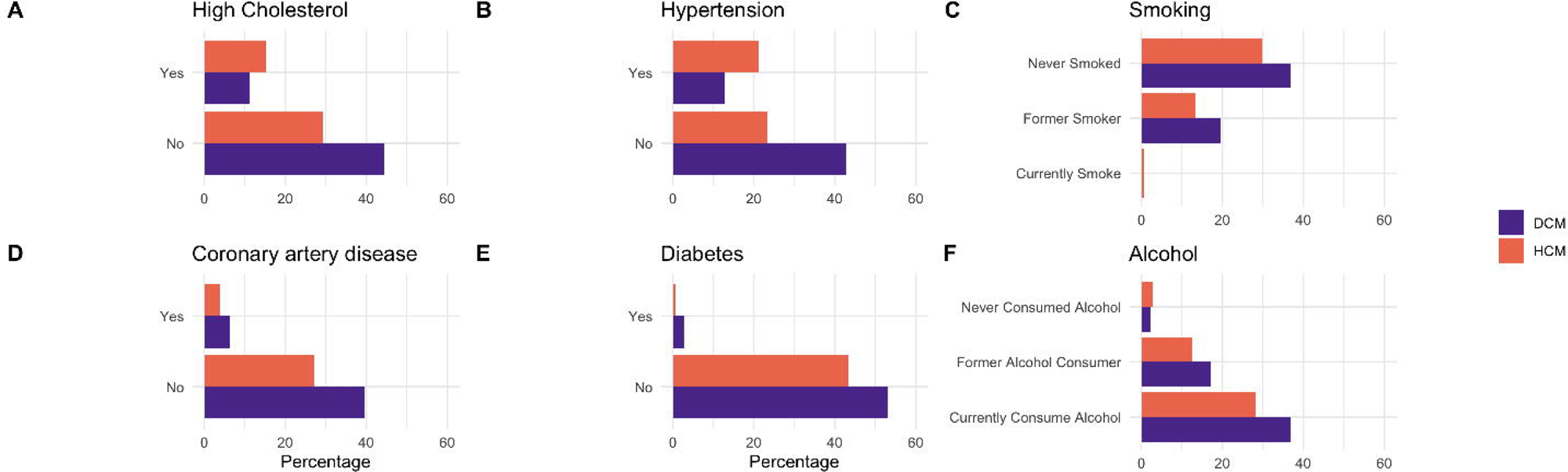
Heart Hive Cardiomyopathy Study participant self-reported risk factor and comorbidity status, alcohol use and smoking status. Unknown/unanswered are not shown– 0.5% did not report diabetes and hypertension status, 1.7 % did not report cholesterol status and 23 % did not report coronary artery disease.

### Comparison with clinic recruited cohort

Compared with a cardiomyopathy cohort traditionally recruited at the Royal Brompton Hospital, a larger proportion of the HH-CM pilot cohort report white European ancestry, 97 % in HH-CM pilot cohort vs 80% in the clinic-recruited cohort (*p = 3.5 × 10^−8^, chi squared test*) and a significantly larger proportion are female – 61% of DCM patients and 53 % of HCM patients were female in the HH-CM pilot cohort compared to 32 % and 28 % respectively in the clinic-ascertained cohort, (DCM *p* = 3.1 × 10⁻^8^, HCM *p = 4.1 x 10^5^*). The two cohorts have similar age ranges (*p* = 0.4, Wilcoxon rank-sum test), although age at diagnosis was younger in the HH-CM pilot group (*p* = 9.9 × 10⁻L, Wilcoxon rank-sum test). The clinic-recruited cohort were recruited close to their diagnosis date – many at diagnosis. Atrial fibrillation (AF) was reported in 51 % of DCM and 46 % of HCM participants in the HH-CM pilot cohort, compared with much lower numbers in the clinic recruited cohort (12.2 % and 16.3 % of DCM and HCM participants) which was recorded at the time of enrolment (DCM *p* = 1.5 × 10⁻^10^, HCM *p = 1.8* × *10^-8^ chi-squared test*), the more granular data collected in the Heart Hive survey shows pre-diagnosis levels of AF are lower (13.2% and 13.1 %). (See supplementary table 1)

### Genetic Burden Testing

The genetic architecture of HH-CM pilot recruited cohorts was consistent with clinic recruited cohorts. There was no significant difference in the total frequency of rare variants in relevant DCM and HCM genes for clinic-recruited versus Heart Hive recruited cohorts. Rare variants were detected in DCM genes in 269/863 (31 %) clinic recruited DCM participants compared to 34/98 (35 %) in the HH-CM pilot cohort (*p= 0.42,* Fisher’s exact tests); and in HCM genes in 153/381 (38 %) clinic-recruited HCM participants compared to 34/76 (45 %) in the HH-CM cohort (*p= 0.30*). Moreover, there was no difference (Fisher’s exact tests) in the distribution of rare variants between genes for each cardiomyopathy type. Variant frequencies and distribution are displayed in Figure 6.

**Fig 6.**
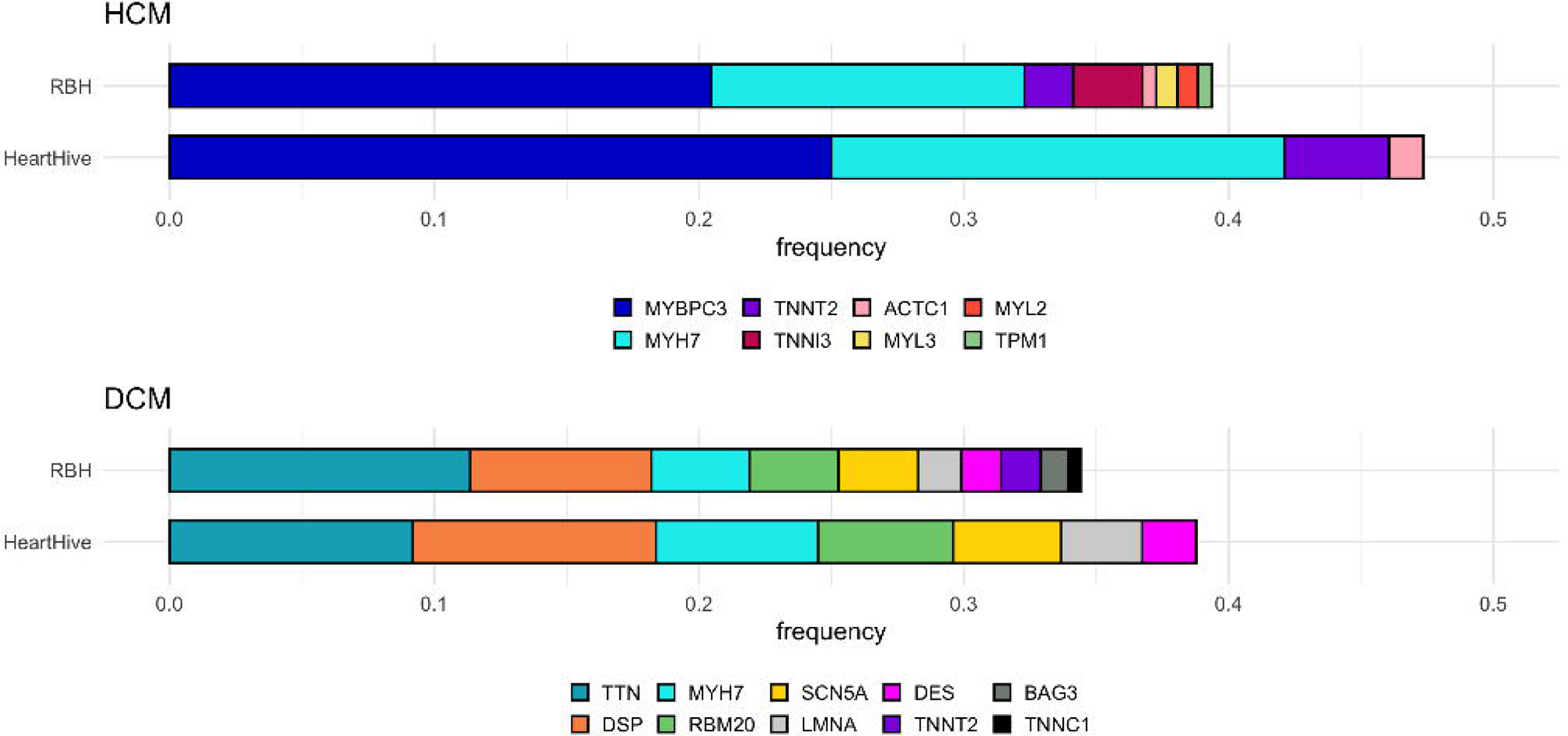
Frequency of rare protein altering variants in genes associated with HCM or DCM found in the Heart Hive cohort vs the clinic recruited cohort from Royal Brompton Cardiovascular Biobank. No differences observed in overall number of variants or by individual genes.

### Returning genetic results in a DTP study

Participants had the option to opt-in to receive research results in relevant cardiomyopathy genes. Results were assessed by accredited clinical scientists. Only variants that were likely to be relevant to participants cardiomyopathy diagnosis (ACMG classified pathogenic or likely pathogenic) were returned to participants via clinicians (Figure 2). 96/98 (98%) DCM and 75/76 (99%) HCM participants, chose to receive personal genetic results. Of these, 16/96 (17%) DCM and 22/75 (29%) HCM were found to have a reportable variant. 59% of the HH-CM cohort self-reported having diagnostic genetic testing at the time of recruitment or after the follow up surveys (n = 45 DCM, 58 HCM). 16% of those with a reported family history of the same cardiomyopathy type had not had diagnostic genetic testing (n= 5 DCM, 5 HCM). In the group without clinical genetic testing, we found reportable variants in 20% of participants (n= 9 DCM, 5 HCM). In those with clinical genetic testing, our panel sequencing identified a reportable variant in 27% of participants, whilst 41% of participants reported a causative variant had been identified. Our pipeline called reportable variants in 5 participants who told us they had a variant of uncertain significance (VUS) or a result classified as ‘other’; 9 participants reported having a causative variant that our pipeline classified as a VUS (or they did not provide variant details, but a VUS was detected); 4 reported variants in regions that were not covered by our panel at that time (FLNC & mitochondrial variants), 1 participant-reported variant was not reported by our pipeline at the time.

## Discussion

The Heart Hive is a valuable way to connect with people across the UK with cardiomyopathy. 78% of participants in the HH-CM study pilot cohort had never previously taken part in cardiovascular research, demonstrating our ability to reach this underserved community. We were successful in recruiting people outside of major cities and specialist ICC centres (44% of participants did not list an ICC centre when asked to name the hospital they received their clinical care), bringing research opportunities to a new set of people with cardiomyopathy, and from a researcher perspective, expanding the reach and diversity of available patient data (Figure 4).

The Heart Hive provides an opportunity to address the underrepresentation of female participants in cardiovascular research. A larger proportion of the HH-CM pilot cohort are female compared to both our clinic recruited cohort and the reported incidence of cardiomyopathy. An enrichment of female participants was expected, as remote⍰Ionly health studies report a median 59-77 % female enrolment, and multiple survey⍰Imethodology papers confirm higher female response rates to web questionnaires [4–6]. Conversely, across conventional interventional studies female patients enrol at roughly half the rate of males, and in many high⍰Iburden areas, for example, cardiovascular disease trials, female participation falls to one⍰Ithird. The evidence suggests persistent structural barriers rather than greater female reluctance to take part in research. [7]

Participation-to-prevalence ratio (PPR) measures how closely study recruitment reflects disease prevalence, a score of 1 is perfect match. Looking specifically at cardiovascular trials, one study found that women constituted only 38 % of participants in 740 cardiovascular trials registered on ClinicalTrials.gov, corresponding to a PPR of 0.81 (classified borderline-low). PPR varied by study type with procedural trials 44 % female vs 78 %% in lifestyle trials [8].

Similarly, male participants consistently make up the majority of published cardiomyopathy cohorts [9]. About 30 % of DCM patients are female [10] giving the Heart Hive a PPR of 1.75 (40 / 76 = 52.6 %); approximately 40 % of HCM patients are female [9, 11, 12] giving the Heart Hive a PPR of 1.63 (60 / 98 = 61.2 %). Our female excess may result in differences compared to typical male dominated cohorts: cardiomyopathies have been shown to have different disease expression in females vs males. In DCM females have been found to have higher rates of adverse heart failure events within two years of diagnosis [9, 13]. In HCM females are more likely to be at an advanced stage of disease at diagnosis compared to males[9], and sex differences have been found to influence risk factors in heart failure [14]. [9] The Heart Hive’s female strong cohort may be a useful resource in addressing questions relating to sex-differences in cardiomyopathy that require more gender balanced cohorts to fully understand. The Heart Hive offers the opportunity to connect researchers to research-willing female participants for trial recruitment to help address this imbalance.

The Heart Hive participants are predominantly of European ancestry groups, with other groups less well represented. We are employing several strategies to capture a wider range of participants, including working with local community groups, and exploring translations and pop-up sessions at community venues. Our participants also tend to have a high level of education and be living in a couple; this may be reflective of the typically older age demographics of a cardiomyopathy cohort. Our participants in the Heart Hive had a very similar age distribution to our clinic-recruited cohort. Although we were initially concerned that relying on an online platform might exclude older individuals and skew participation toward a younger demographic, this did not appear to be the case. Despite the similar age range for the Heart Hive and clinic recruited cohorts, our clinic recruited cohort signed up much closer to their diagnosis date and some differences between the cohorts may reflect this. For example, many of our clinic cohort were recruited at attendance for their first MRI which raises the question of whether the clinic cohort could be depleted of known arrythmias on their baseline assessment.

At the individual patient level, self-reported genetic findings were not precise, and a broad range of patient literacy was observed. Some participants could provide full genetic variant details and others could recall a positive or negative result but not gene names. Variants detected in our research bioinformatic pipeline had not always been identified by participants or had been categorised differently (e.g. our pipeline classified a variant as VUS and participants described as causative). It is difficult to know whether the complexities of genetic results are always well understood by patients, which cardiomyopathy related genes have been included in historical panels or single gene assays, or if discrepancy arises simply because variant calling pipelines have improved over time. Equally, clinical teams may have access to segregation data that our research team did not. The mixed concordance of participant-reported genetic results with our in-house research results highlights the absolute necessity of carrying out our own sequencing to fully understand the genetic architecture of the cohort.

Our genetic analyses show the cohort recruited through the Heart Hive are genetically equivalent to traditionally recruited cohorts, suggesting that self-reported diagnoses are accurate and reliable, and supporting the direct to participant recruitment model as an effective and scalable option.

In the group of participants who did not report receiving clinical genetic testing we found clinically actionable likely pathogenic or pathogenic variants in 18% which suggests there is an unmet need for timely genetic testing for cardiomyopathy patients and that there may be disparity in access to genetic testing within the UK. This also highlights the utility and feasibility of mail-out genetic testing, for example to assess eligibility for clinical trials.

The Heart Hive demonstrated a high level of patient engagement and retention. Attrition is a well-recognised threat to statistical power in clinical research [15–17] and tends to be even higher in technology⍰Imediated research: a 36⍰Itrial meta⍰Ianalysis of smartphone⍰Ibased diabetes interventions found an overall dropout of 29.6 % (95 % CI 25 - 34 %) [18], while a systematic review spanning multiple chronic⍰Idisease mobile-app based studies estimated a pooled attrition of 43 % (95 % CI 29–57 %) [19]. Observational cohorts are only modestly better, with a 143⍰Icohort meta⍰Ianalysis reporting a mean retention of 73.5 % (SD 20.1 %), equivalent to 26 % attrition [20], and fully remote cardiovascular e⍰Icohorts such as the electronic Framingham Heart Study retaining just 59 % of participants by three months [21]. By contrast, the Heart Hive Cardiomyopathy Study achieved an 86 % saliva⍰Ikit return and 83 % follow⍰Iup survey completion, yielding an overall attrition of only 14–17 %. These figures fall well below the 20 % threshold commonly regarded as the point at which attrition materially threatens validity and markedly outperform both on⍰Isite clinical trials and comparable digital cohorts. We attribute this high engagement to the platform’s patient⍰Iled co⍰Idesign, automated yet personalised prompts, low participant burden (entirely home⍰Ibased procedures), and the return of individual study results, all of which align with retention⍰Ienhancing strategies identified in methodological reviews. Nonetheless, follow⍰Iup remains shorter than in decades⍰Ilong legacy cohorts, and the self⍰Iselected, digitally literate sample may limit external generalisability; ongoing monitoring will be necessary to confirm the sustainability of these early retention gains.

Research participants report that they like to hear the outcomes of research at the end of the study [22]. The Heart Hive functions as an effective and efficient way to directly return study updates and outcomes to the community through email, the dashboard, blogs, and social media channels. During the COVID-19 pandemic we were able to mount an agile response and rapidly gauge the impact on cardiomyopathy patients by leveraging a turnkey registry and contact portal, coupled with a pre-registered research-willing community. We rapidly deployed a survey to understand how people with cardiomyopathy experienced healthcare during the COVID-19 pandemic, and how the pandemic, lockdowns, and changes to delivery of routine cardiomyopathy care impacted patients’ wellbeing [23].

The Heart Hive aligns closely with patient-identified priorities in cardiomyopathy research. Recent priority-setting exercises, such as those led by Cardiomyopathy UK in partnership with the James Lind Alliance, have underscored the need to better understand disease progression, improve access to genetic testing, and promote more inclusive research participation [24]. Patients, carers, and clinicians jointly identified these as key areas to guide future research and policy. The Heart Hive directly addresses these priorities by enabling inclusive, patient-centred participation, supporting longitudinal tracking of disease progression, and expanding access to genetic testing through remote sample collection.

Patient advocacy organisations such as Cardiomyopathy UK have emphasised that patients often find participation in research empowering and value the opportunity to contribute to advances in care. By enabling individuals to proactively engage with studies through remote consent, health data sharing, and return of results, The Heart Hive directly supports this agenda, transforming patients from passive recipients of care into active partners in research.

The accumulated experience of several mature direct-to-patient (DTP) platforms shows that remotely collected data can equal or even outperform clinic-based cohorts when properly validated. Fox Insight (≈ 54 000 registrants) verified 203 self-declared Parkinson’s disease diagnoses by structured video consultations with movement-disorder specialists and found “very good” agreement[25]. The Swiss Multiple Sclerosis Registry (≈ 4 000 participants) demonstrated substantial reliability after linking survey responses to compulsory health-insurer claims, for both disease-modifying-therapy exposure and MS subtype [26]. Count Me In (>100,000 enrolees) is a pan-cancer, patient-partnered initiative that invites any U.S. or Canadian patient with any malignancy to enrol online and share tumours, blood and medical records; The Count Me In Angiosarcoma Project harnessed an exceptionally active on-line community to enrol 338 patients, exceeding the cancer’s annual US incidence in 18 months, and its first genomic release identified recurrent mutations that now inform therapeutic studies [27]. The UK GLAD Study has recruited > 40 000 people with lifetime anxiety or depression entirely on-line; although it does not cross-check medical records, diagnoses are assigned with validated instruments (CIDI-SF, PHQ-9, GAD-7), giving acceptable sensitivity and specificity for DSM disorders [28].

Together with Heart Hive, these exemplars rebut the notion that remote, participant-led recruitment compromises diagnostic validity and provide a benchmark for other DTP platforms. The Heart Hive bridges the gap between population biobanks and ultra-rare disease registries, offering a scalable, trial-ready model tailored to cardiomyopathies. By combining remote genomics, real-time outcome capture, and rapid, patient-driven adaptability, it enables inclusive, genotype-directed research without clinic bottlenecks—setting a blueprint for future rare disease studies.

Our re-contactable design enables rapid identification of genotype-eligible participants for mechanistic studies and precision-therapy trials. Genetically, cardiomyopathy is dominated by highly penetrant single-gene variants (>60 causal genes), making it directly actionable. In addition to the first PKP2 AAV gene-replacement trials now open (TN-401, RP-A601, LX2020) [29–31], relevant programme examples include MYBPC3-HCM gene replacement (TN-201) [32], LAMP2 (Danon disease) gene therapy (RP-A501) [33, 34], and BAG3-DCM gene replacement (RP-A701) [35]. Heart Hive’s genomics-first, re-contactable architecture is designed to match patients to such studies quickly and at scale.

The Heart Hive registry has always welcomed participants with any cardiomyopathy, as well as those with inherited risk. The Cardiomyopathy Study component—initially focused on HCM and DCM—has now broadened to include additional phenotypes (e.g., Takotsubo cardiomyopathy and ARVC) and is transitioning from panel testing to whole-genome sequencing to capture both rare and common variant effects.

Heart Hive is available as a platform for investigator-initiated cardiovascular research in the UK and internationally. To date it has supported patient surveys, observational cohort work (including registry-based designs), and targeted recruitment to interventional trials; a current directory of hosted studies is available at thehearthive.org/research.

Our participant community is the engine of Heart Hive. Through trusted partnerships with patient groups and embedded PPIE, participants shape study priorities and design, co-design questionnaires, and keep our methods grounded in lived experience. This trust translates into sustained engagement, rapid and ethical re-contact for genotype-eligible studies, and a reach that extends beyond tertiary clinics. In Heart Hive, participants are partners rather than subjects—making the research faster, fairer, and more likely to influence care.

At its heart, this is research with, rather than on, participants. The Heart Hive’s participant community are keen to be involved with research opportunities. With mail-out genomics, high retention, and a fully digital, low-barrier design, Heart Hive enables trial-ready, genotype-informed studies while keeping patients at the centre. This partnership model delivers clinic-grade data today and a faster path to better treatments tomorrow.

Box 1 – survey to assess for ischaemic cardiomyopathy

## Data Availability

All data produced in the present study are available upon reasonable request to the authors

## Acknowledgement

We extend our sincere thanks to all Heart Hive participants who signed up to the portal and consented to take part in this research. Our community of participants has been instrumental in shaping the study through their feedback and engagement at every stage. This work has been supported by Cardiomyopathy UK, whose guidance, advice, and commitment to patient-led research have been invaluable to the success of the project. We also thank Crowdfunder UK for their support during the fundraising and pre-launch stages, and the many individual donors who contributed. Finally, we acknowledge Broad Clinical Labs for partnering with us to rebuild the Heart Hive on the Juniper data donation platform.

For the purpose of open access, the authors have applied a Creative Commons Attribution (CC BY) licence to any Author Accepted Manuscript version arising. The views expressed in this work are those of the authors and not necessarily those of the funders.

---

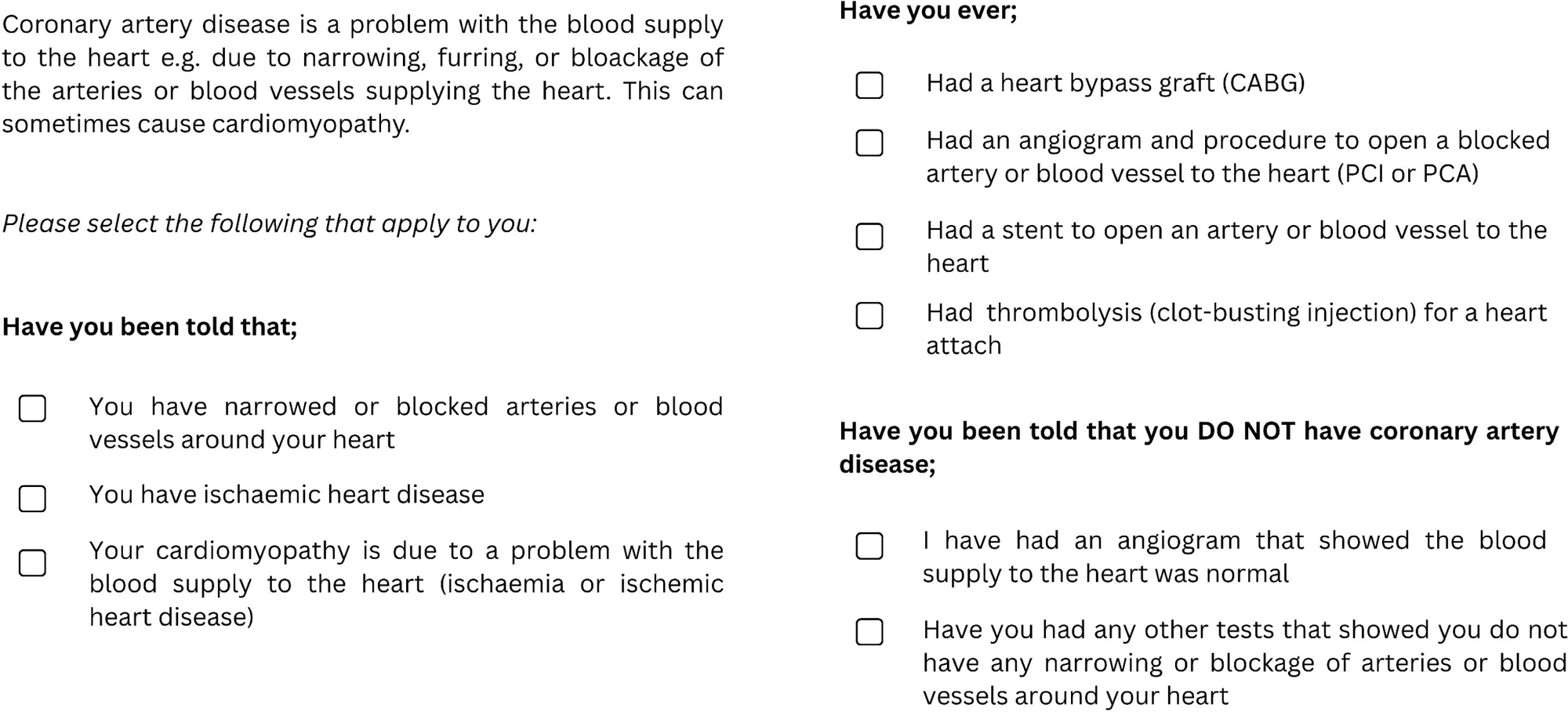

