## Supplemental Table 1 for "The Heart Hive: direct-to-participant recruitment provides an effective, scalable route to identify engaged participants for cardiomyopathy research"

**Sociodemographic, behaviour, and selected clinical history (descriptive only)**

| **Variable** | **Heart Hive (HH)** | | **RBH clinic cohorts** | | **National surveys** | | | **UK register ‑based** |
| --- | --- | --- | --- | --- | --- | --- | --- | --- |
|  | **DCM (n=98)** | **HCM (n=76)** | **DCM (n=863)** | **HCM (n=381)** | **England** | **Wales** | **Scotland** | **(where applicable)** |
| **Age at recruitment, median (IQR) (y)** | 56.5 (45-66) | 58.5 (50-66) | 56 (45-65) | 57 (46-66) | — | — | — | — |
| **Age at diagnosis, median (IQR) (y)** | 48.5 (42-59) | 49 (41-59.5) | 53 (43 – 62) | 55 (43-65) | — | — | — | — |
| **Population age median (IQR)** | — | — | — | — | 48 (34-63) | 51 (34-66) | 50 (34-64) |  |
| **Sex: female** | 61.20% | 52.60% | 32.40% | 27.80% | 51.% | 51% | 51% | — |
| **White (census ethnic group category) ^a^** | 96.90% | 98.70% | 83.80% | 75.30% | 81.0% | 93.8% | 92.8% | — |
| **Asian (census category)** | — | — | — | — | 9.6% | 2.9% | 3.4% | — |
| **Black (census category)** | — | — | — | — | 4.2% | 0.9% | 0.12% | — |
| **Graduate degree**  **(Level 4+ / SCQF 7–12) ^b^** | 53% (HH overall) | | — | — | 27.60% | 25.9% | 32.5% | — |
| **A level or equivalent**  **(Level 3 / SCQF 6)** | 32% (HH overall) | | — | — | 13.78% | 14.18% | 11.1% | — |
| **GCSE or equivalent**  **(Level 2 / SCQF 5)** | 12% (HH overall) | | — | — | 10.84% | 11.82% | 18.9% | — |
| **Lives in a couple household** | 72.4% | 76.3% | — | — | 61% | 60% | 40% | — |
| **Congenital heart disease (history)** | 4.0% | 9.5% | — | — | — | — | — | 2% |
| **Stroke/TIA (history)** | 2.0% | 5.4% | — | — | 1.8% | 2.2% | 2.3% | 1.90% |
| **Heart failure (history)** | — | — | — | — | 1% | 1.1% | 0.8% | 1.00% |
| **Coronary artery disease (history)** | 11.2% | 9.2% | — | — | 3% | 3.6 | 3.9% | 3.1% |
| **Atrial fibrillation (ever)** | 50.7% | 46.0% | 21.2% | 16.3% | 2.1% | 2.4% | 1.9% | 2.1% |
| **Atrial fibrillation before cardiomyopathy diagnosis** | 13.2% | 13.1% | — | — | — | — | — | — |
| **Family history HCM/DCM (known)** | 33.6% | 43.4% | 11.8% | 22.0% | — | — | — | — |
| **Diabetes (ever/diagnosed)** | 5.1% | 1.3% | 13.8% | 9.7% | 7.7% | 8.2% | 7% | — |
| **History of hypertension (ever)** | 23.1% | 47.3% | 30.0% | 40.4% | 14.8% | 16% | 13% | 15% |
| treated & controlled | 20.0% | 38.2% | — | — |  |  |  | — |
| untreated / uncontrolled | 3.1% | 9.1% | — | — | — | — | — | — |
| **History of high cholesterol (any)** | 20.0% | 34.2% | — | — | 53% | — | — | — |
| **treated & controlled** | 12.6% | 23.7% | — | — | — | — | — | — |
| **untreated / uncontrolled** | 7.4% | 10.5% | — | — | — | — | — | — |
| **Valve disease** | 22.4% | 32.9% | — | — | — | — | — | — |
| **Chronic kidney disease (history)** | 5.1% | 1.3% | — | — | 4.4% | — | — | — |
| **Implanted cardiac device** | 44.9% | 27.6% | 32.9% | 10.5% | — | — | — | — |
| **Obstructive HCM (LVOT)** | — | 36.8% | — | 19.4% | — | — | — | — |
| **History of chemotherapy** | 2.0% | 0.0% | 4.6% | — | — | — | — | — |
| **History of smoking (ever)** | 34.7% | 31.6% | 40.5% | 40.7% | 38% | 38% | 38% | — |
| **Current smoker** | 0% | 1% | 9.2% | 8.1% | 13% | 14% | 15% | — |
| **Ex ‑smoker** | 34.7% | 30.2% | — | — | 25% | 24% | 25% | — |
| **Current alcohol drinker (status, HH/RBH)** | 65.3% | 64.5% | 38.9% | 37.8% | — | — | — | — |
| **Current drinker (national, last 12 months) ^c^** | — | — | — | — | 81% | 83% | 80% | — |

**General comparability note:** HH values are self reported; RBH values are clinically recorded; national figures combine survey measured and register based estimates with differing sampling frames and definitions. The table is descriptive only; no statistical comparisons are presented.

**Footnotes**

^a^ Ethnicity rows use the national census *ethnic group* categories (e.g., “White”, “Asian”, “Black”), not genetic or self defined ancestry. England & Wales figures are from the 2021 Census; Scotland figures are from Scotland’s Census 2022. Category labels differ slightly between nations; values are shown as published.

^b^ Education categories are aligned to UK qualification levels: Graduate degree = Level 4+ (degree or higher; SCQF 7–12); A level or equivalent = Level 3 (SCQF 6); GCSE or equivalent = Level 2 (SCQF 5). HH overall split: 53% graduate degree, 32% A level/equivalent, 12% GCSE/equivalent (values represent highest qualification; residual may reflect other/unknown categories).

^c^ Status, last 12 months – proportion reporting any alcohol use in the previous year.
